# High levels of mild to moderate depression among men who have sex with men and transgender women in Lima, Peru: implications for integrated depression and HIV care

**DOI:** 10.1101/2020.01.07.20016808

**Authors:** Jerome T. Galea, Stephanie Marhefka, Segundo R. León, Guitele Rahill, Elena Cyrus, Hugo Sánchez, Zhiwei Zhang, Brandon Brown

## Abstract

Depression disproportionally affects people at risk of acquiring or living with HIV and is associated with worse health outcomes; however, depression care is not routinely integrated with HIV prevention and treatment services. Selection of the best depression intervention(s) for integration depends both on the prevalence and severity of depression among potential users. To inform depression care integration in a community-based setting in Lima, Peru, we retrospectively analyzed routinely collected depression screening data from men who have sex with men and transgender women seeking HIV prevention and care services (N=185). Depression was screened for using the Patient Health Questionnaire-9. Prevalence of any depression (PHQ-9 ≥5) was 42% and was significantly associated with the last sexual partner being “casual” (p=0.01). Most (81%) depressive symptoms were mild to moderate (≥5 PHQ-9 ≤14). Integrating depression care with HIV prevention and treatment services in Peru should begin by implementing interventions targeting mild to moderate depression.

## INTRODUCTION

Globally, depression among people at risk of acquiring HIV and people living with HIV (PLWH) significantly worsens health outcomes and requires urgent action^[1-6]^. Untreated depression is associated with behaviors that increase HIV acquisition risk including substance use, condomless sexual intercourse, and attrition from HIV prevention programs^[4,7-11]^. For PLWH, depression doubles the risk of mortality^[12]^, lowers uptake and adherence to medical care^[13,14]^, and is associated with viral load increases^[15]^. Evidence supports the benefits of depression care for these populations ^[16-19]^ and public policy—including the Joint United Nations Programme on HIV/AIDS (UNAIDS) and the U.S.’s new “Ending the Epidemic: A Plan for America”—considers mental health service integration with HIV prevention and treatment services essential to curtailing the HIV epidemic^[20-22]^. Despite the evidence base and political support, however, depression screening and care is not routinely integrated with HIV prevention and treatment services, constituting a mental health and HIV service “gap”^[21]^.

A major barrier to integrated depression and HIV prevention and treatment stems from a traditional paradigm of mental health care delivery models relying on highly skilled mental health workers (e.g., psychiatrists, psychiatric nurses, and trained psychosocial care providers), and a simultaneous global shortage of these professionals. In low- and middle-income countries (LMICs), for example, there is on average less than 1 psychiatrist for every 100,000 persons^[23]^. Even in high-income settings like the U.S., a severe shortage of mental health professionals has been reported, especially in rural settings^[24]^. At the same time, LMICs are also home to >90% of people living with HIV^[25]^. Integrated depression/HIV care models that rely solely on skilled mental health professionals are therefore unlikely to be feasible in many settings.

Promising alternatives to the traditional mental health service models involve non-specialist health workers (NSHWs)—individuals that have not been formally trained in mental health care—to deliver depression care instead of specialized mental health professionals^[26]^. In these models, NSHWs learn to screen patients and deliver evidence-based, low-intensity, non-pharmacological, psychological treatment for mild to moderate depression, referring patients with serious depression to mental health professionals. The interventions that NSHWs deliver are called “low-intensity” because they use fewer resources than more traditional psychological interventions and can be delivered by people with no formal training in mental health. Growing from the much larger field of Global Mental Health^[27]^, which has demonstrated success in NSHW-delivered, low-intensity depression interventions to diverse populations in LMICs^[28]^, NSHWs^29^ could be trained to screen and treat depression. Indeed, NSHWs already play a critical role in delivering HIV prevention and care services in LMICs^[29]^. In such a “stepped-care” model, the intensity of the intervention is matched to the patient’s level of depression severity. For example, patients screening for low or moderate levels of depression would be offered self-help and/or NSHW-delivered depression care while more specialized psychotherapies and antidepressant medications (and the specialized staff required to deliver them) would be reserved for patients with more severe depression symptomology^[30-34]^.

Peru, a middle-income country, continues to experience an unabated HIV epidemic that is highly concentrated in men who have sex with men (MSM) and transgender women (TW), with prevalence rates of 13%^[35,36]^ and 30%^[37]^, respectively, compared to a general adult population HIV prevalence rate of 0.01-0.03%^[38]^. Though the Peruvian Ministry of Health has made significant strides towards increasing access to mental health services nationally over the past five years^[39]^, and depression is recognized in the country’s technical guide for treating HIV as a risk factor for poorer antiretroviral therapy (ART) adherence^[40]^, evidence-based depression screening and care is not routinely integrated with HIV prevention and treatment services. Nonetheless, some limited data on depression prevalence and severity in Peruvian populations at risk of HIV acquisition and PLWH begin to characterize the problem.

One study by Maldonado et al^[41]^ measured the frequency of a major depressive episode and related factors among recipients of ART in a public hospital in Lima (N=205). Depression was measured using a locally validated, 5-item version of Center for Epidemiologic Studies Depression scale (CES-D)^[42]^, which has a point range of 0-15; higher scores are indicative of more depressive symptomology. In this study, the frequency of major depressive episode, defined by a cut-off score of 6, was 27.8% among a sample that was mostly male (N=139) and among whom 65.9% self-identified as heterosexual. The authors concluded that the prevalence of a major depressive episode among study participants was higher than the prevalence in the general population but not associated with ART adherence.

In another study, Ferro et al found depressive symptoms prevalent in 44.5% of a study population comprised of Peruvian MSM and TW who were diagnosed with HIV for >1 year (N=302)^[43]^. In the study, which was designed primarily to elucidate the effect of alcohol use disorders on ART adherence, depression was assessed using the 10-item version of the CES-D^[44]^; a cutoff of score of >7 was used for moderate to severe depressive symptoms. While depression was not independently associated with poor ART adherence, it was highly correlated with problem alcohol use which was associated with ART non-adherence, and the authors recommend dual screening and treatment for both disorders.

A third study, though not exclusively conducted in Peru, sheds light on depression among populations at risk for HIV. The iPrEx study (which demonstrated the efficacy of daily oral Truvada® to prevent HIV acquisition, i.e “PrEP”, among MSM and TW^[45]^), also assessed depressive symptoms among study participants using the 20-item version of the CES-D. In this version, scores range from 0-60, again with higher scores indicative of more depressive symptomology; scores of ≥16 have been regarded as clinically significant^[46]^. In the iPrEx depression analysis, among all (HIV-negative) participants with at least one CES-D score (N=2,131, of whom 1,172 participants were from Peru), the mean CES-D score was 14 (range 0-55), with those in the highest quartile (CES-D ≥20) most likely to report condomless receptive anal intercourse during the previous 3 months^[47]^. Depression was not associated with PrEP use, though, notably, half of all participants in iPrEx had CES-D scores of ≥16 over the course of the study.

While the preceding studies help characterize depression among Peruvian MSM and TW living with or at risk of HIV and each indicates the necessity for depression care for these populations, important questions remain regarding the type(s) of depression interventions that could target these groups. As previously noted, relative depression severity is especially important to understand during the development of integrated depression and HIV care models because it informs both the interventions used (e.g., psychological and/or pharmacological) and the level of staff expertise required (e.g., psychiatrist, other mental health specialists, NSHW, layperson) needed to deliver the intervention. The previous studies found that higher levels of depression were common among the study populations; however, this information is insufficient to inform depression intervention choices in the context of integrated depression/HIV care. Here, we report depression prevalence and severity among attendees at a community-based clinic serving MSM and TW at risk for or living with HIV in Lima, Peru, information used to inform a process of depression care integration in this setting.

## METHODS

### Participants, Procedures

We conducted a retrospective analysis of anonymized, existing clinic data from individuals seeking HIV testing between August 2017 and December 2018 at Epicentro Salud, a non-profit organization that provides a range of services serving the LGBTQ population in Lima, Peru including a sexual health clinic. Social and psychological support services are provided in the form of support groups and individual counseling delivered by bachelors-level psychologists while the sexual health clinic provides free or low-cost sexually transmitted infection (STI) testing and treatment and HIV testing with linkage to external PrEP or treatment providers. Epicentro’s clientele is characterized by being mostly *mestizo/a*, 20-35 years of age and predominantly men who self-identify as gay and some TW. Clients seeking sexual health services are attended to by a physician who assesses their risk for HIV (if HIV status is unknown or if the previous HIV test was negative) and other STIs using an in-house sexual health and risk assessment survey which includes depression screening (see Measures below).

HIV testing is conducted using the Determine HIV-1/2 Ag/Ab Combo rapid test (ALERE Healthcare, S.L.U). Clients testing positive for HIV are referred to the national HIV program for free confirmatory testing and treatment and invited to participate in a weekly support group offered at Epicentro for PLWH. Symptomatic STIs are treated empirically or further testing and/or referrals are offered. Clients screening positive for mild or greater depression are provided brief psychoeducation and referred to existing community-based mental health services for clinical evaluation. Those with moderately severe or greater depression and/or any suicidal ideation are attended to by a staff psychologist (HS) at Epicentro and receive additional evaluation, referrals, and accompaniment to specialized mental health services.

### Measures

The physician-administered, in-house sexual health and risk assessment is comprised of 21 questions that collect information on age; motive for clinic visit (routine check-up or in response to a known sexual risk behavior); sexual and gender self-identification (homosexual or gay, bisexual, heterosexual, transgender); presence of STI symptoms; most recent sexual partner (number of days since last sexual intercourse); partner type (stable/regular, casual, “one-nighter”; condom use; and, alcohol and/or drug use. Depression screening is conducted using the Patient Health Questionnaire-9 (PHQ-9), originally designed and validated for depression screening in primary care/clinical care settings^[48]^, with a Spanish version validated for use in Peru in clinical settings^[49]^. Standard cut-off scores for the PHQ-9 were used: minimal/no depression (PHQ-9 = 0-4); mild depression (PHQ-9 = 5-9); moderate depression (PHQ-9 = 10-14); moderately severe depression (PHQ-9 = 15-19); and severe depression (PHQ-9 = 20-27)^[48]^.

### Data Analysis

Overall depression prevalence and severity were calculated and reported according to the PHQ-9 cutoff scores. To assess the relationship between depression, sexual health, and sexual behavior, we conducted statistical tests of bivariate association considering one characteristic at a time. Categorical variables were tested using Pearson’s chi-square test. Continuous variables were tested using a two-sample t-test (for apparently normally distributed data) and Wilcoxon’s rank-sum test (for clearly skewed data). All tests were conducted using R version 3.5.2 (2018).

### Statement on Human Subjects

The research ethics committees for Epicentro in Peru and the University of South Florida designate retrospective studies utilizing existing data with no personal identifiers as non-human subjects research.

## RESULTS

### Study Population

Data from 185 MSM and TW who presented to Epicentro seeking services from the sexual health clinic during the reporting period were included in the analysis. Median patient age was 27 years (range 17-58), and 58% (107/185) were seeking routine HIV and/or STI testing (i.e., not in response to a specific sexual risk event, but as a regular sexual health check-up). Eighty-five percent (158/185) of clinic attendees self-identified as homosexual or gay; 9% (17/185) as bisexual; 3% (5/185) as heterosexual; and 3% (5/185) as TW. Thirteen percent (24/181) reported living with HIV, of which 83% (20/24) were new infections detected during the clinic visit (see Table I).

**Table I:** Descriptive characteristics of Peruvian MSM and TW seeking HIV services in Lima, Peru (N = 185)

| Characteristics | Total<sup>*</sup><br>n (%) | PHQ-9<sup>†</sup> < 5 | PHQ-9<sup>†</sup> ≥ 5 | P value<sup>§§</sup> |
| --- | --- | --- | --- | --- |
|  |  | n (%) | n (%) |  |
| **Age, years** | 27 (23-33)<sup>‡</sup> | 27 (23-32)<sup>‡</sup> | 27 (23-34)<sup>‡</sup> | 0.70 |
| **Clinic visit motive** |  |  |  | 0.85 |
| Routine check-up | 107 (58) | 63 (59) | 44 (41) |  |
| Recent known sexual risk behavior | 78 (42) | 44 (56) | 34 (44) |  |
| **Self-identification<sup>§</sup>** |  |  |  | 0.83 |
| Homosexual or gay | 158 (85) | 93 (59) | 65 (41) |  |
| Bisexual | 17 (9) | 9 (53) | 8 (47) |  |
| Heterosexual | 5 (3) | 3 (60) | 2 (40) |  |
| Transgender woman | 5 (3) | 2 (40) | 3 (60) |  |
| **HIV result** |  |  |  | 0.48 |
| Positive | 24 (13) | 16 (67) | 8 (33) |  |
| Negative | 157 (87) | 89 (57) | 68 (43) |  |
| **STI symptoms present<sup>¶</sup>** |  |  |  | 0.70 |
| Yes | 47 (26) | 26 (55) | 21 (45) |  |
| No | 135 (74) | 81 (60) | 54 (40) |  |
| **Most recent sexual intercourse** |  |  |  |  |
| Days ago | 14 (6-30)<sup>‡</sup> | 7 (6-30)<sup>‡</sup> | 18 (7-56)<sup>‡</sup> | 0.07 |
| **Partner type** |  |  |  | **0.01** |
| Stable/regular partner | 68 (37) | 48 (71) | 20 (29) |  |
| Casual or 'one-nighter' | 117 (63) | 59 (50) | 58 (50) |  |
| **Condom used** |  |  |  | 0.64 |
| Yes | 106 (58) | 64 (60) | 42 (40) |  |
| No | 77 (42) | 43 (56) | 34 (44) |  |
| **Alcohol and/or drug use<sup>\*\*</sup>** |  |  |  | 0.38 |
| Yes | 34 (18) | 17 (50) | 17 (50) |  |
| No | 150 (82) | 90 (60) | 60 (40) |  |

\* Due to missing data, not all variables sum to the total N.
† PHQ-9: Patient Health Questionnaire
‡ median (IQR)
§ Though not mutually exclusive categories, clinic attendees self-identified as belonging to only one of these groups.
¶ STI symptoms included: burning while urinating; perianal/genital warts; ulcers on penis and/or in anus; penile and/or anal discharge; and genital rash.
\*\* Drugs included: marijuana, cocaine, poppers, and other.
§§ P-values are based on Pearson's chi-square test for all qualitative characteristics, two-sample t-test for age, and Wilcoxon's rank-sum test for "days ago" (due to high skewness and extreme values in data).

### Depression Prevalence and Severity

Overall depression prevalence (PHQ-9 ≥5) was 42% (78/185), of which 81% (66/78) of participants had depression scores that clustered in the mild to moderate severity range (5≤ PHQ-9 ≤14). Depression severity per the 6 PHQ-9 cutoff scores was: minimal/none (PHQ-9 = 0-4): 58% (107/185); mild (PHQ-9 = 5-9): 23% (43/185); moderate (PHQ-9 = 10-14): 12% (23/185); moderately severe (PHQ-9 = 15-19): 5% (9/185); and severe (PHQ-9 = 20-27): 2% (3/185) (see Figure 1).

**Figure 1:**
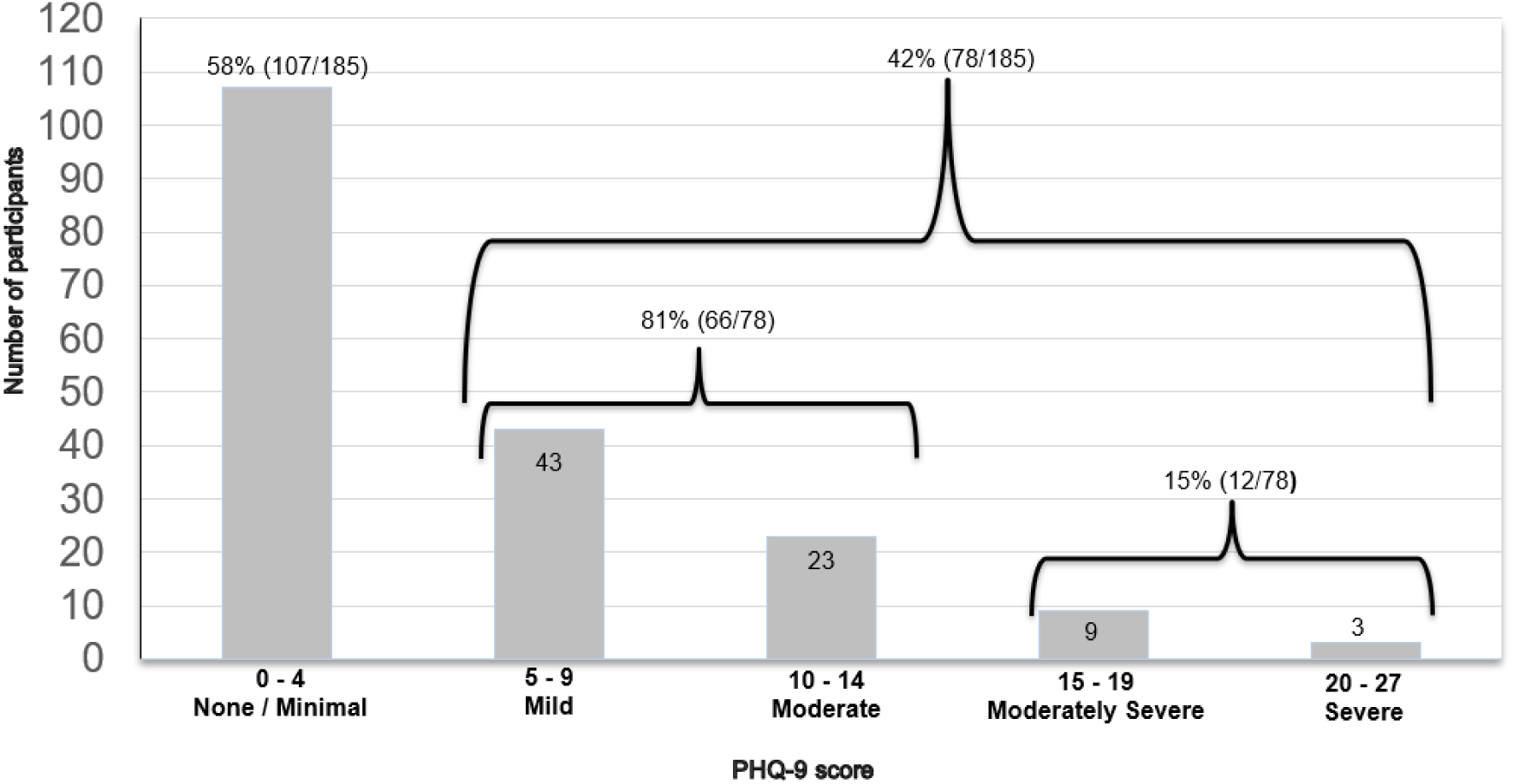
Distribution of the prevalence and severity of depression among Peruvian MSM and TW (N = 185)

### Bivariate Analysis

In the bivariate analysis, screening positive for mild or greater depression (PHQ-9 ≥5) was significantly associated with the most recent sexual partner being a casual or “one nighter” partner versus a stable/regular partner (p=0.01). Also, there was a marginally significant relationship (p=0.07) of clinic attendees with mild or greater depression to go longer without sexual intercourse than those with minimal/no depression (see Table I). No association between depression and HIV infection was observed.

## DISCUSSION

In this retrospective analysis from MSM and TW seeking sexual health services at a community-based health center, greater than minimal depression (PHQ-9 ≥5) was detected in 42% of clinic attendees. In contrast, the prevalence of any depression in this sample was 15 times greater than the 2.8% point prevalence of depression among adult Peruvians in metropolitan Lima^[50]^; however, the use of different instruments, variability of settings (i.e., clinic in our study versus random household survey), and populations limits direct comparison.

For the same reasons, the depression scores found in our study are difficult to directly compare with the previously discussed studies measuring depression among Peruvian MSM and TW^[41,43,47]^; however, ours and the previous studies confirm that depressive symptoms are common in these populations regardless of HIV status. To the best of our knowledge, this is the first report of a community center in Peru conducting routine depression screening as a standard component of sexual health services for MSM and TW, regardless of HIV status.

We found that depression was significantly associated with the last sexual partner being casual (versus a stable or regular partner). While the relationship between mood and sexual behavior is complex, a possible interpretation of this finding is that participants with depressive symptomology sought to self-remedy a negative mood state—albeit in a transitory manner—by seeking contact with others^[51]^.

A central finding in this study was that most (81%) depression scores fell in the mild to moderate range of severity (5≤ PHQ-9 ≤14), which carries important implications. Moderate levels of depression—but neither low nor high depression levels—have been associated with increased sexual risk-taking in both MSM at risk of HIV acquisition^[52]^ and MSM living with HIV^[53]^. Though absent in the present study, if future research bears out this association in Peruvian MSM and TW, efforts towards integrating depression care with HIV prevention and treatment programs should target moderate depression because of its unique association with HIV risk and because it may be the most prevalent of depression severities. Importantly, we do not advocate the non-prioritization of severe depression care for these populations. Instead, we recommend that depression screening in HIV service settings allows for meaningful differentiation of depression severity (as opposed to only “some depressive symptomology” versus “no depression”) and that the treatment offered is commensurate to the detected depression severity.

Prioritizing treatment for moderate depression in HIV service settings, however, presents challenges but also opportunities. Because of the previously described shortage of specialized mental health workers, when mental health services are provided in resource-constrained environments, the most severe cases are prioritized even though they may be limited in number when compared to those with less severe mental health morbidities. And yet, in the context of depression care integration with HIV services, those with moderate depression may be the most frequent in number and are the most likely to engage in HIV risk transmission behaviors^[52,53]^. Finding the resources to expand care beyond the most acute cases is the primary barrier facing mental health service expansion globally. Fortunately, this is precisely where NSHW-delivered, evidence-based depression interventions are ideal because they allow the expansion of depression care to more people without sacrificing necessary mental health professional intervention with more severe cases. Many LMICs are already integrating depression care delivered by people without previous mental health training in general population, community-based settings^[54]^. The challenge is to expand these already functioning programs into the priority area of HIV prevention and treatment.

There is also an opportunity to capitalize on the broader Global Mental Health mandate to increase mental health services for all people in need by integrating existing, NSHW-delivered depression interventions into HIV prevention and treatment programs rather than designing novel interventions. Over 90 LMICs^[55]^ are expanding depression care accessibility using low-intensity psychological interventions disseminated by the World Health Organization’s (WHO) Mental Health Gap Programme (mhGAP)^[56]^. These countries may find it easier and more economical to expand an existing WHO low-intensity depression intervention into the HIV service delivery platform, capitalizing on a “ready workforce” of NSHW already responsible for delivering most HIV prevention and treatment in LMICs^[21]^. While these low-intensity interventions are not the panacea for all types of depression and may require adaptation for use in HIV service settings, their use could be a significant first step towards concretely addressing the problem of depression among those at risk for or living with HIV in a scalable fashion.

### Limitations

Because this was a retrospective, cross-sectional analysis of existing data from a single clinic using forms not designed as research instruments, many important factors which could impact depression in MSM and TW were not addressed. These include a history of violence and trauma; stigma; disclosure of sexual orientation, gender identity, and HIV status; income; more detailed information regarding alcohol and drug use; and adherence to HIV medications for PLWH. A single depression score can only reflect the time frame at which it was administered (for the PHQ-9, this was the previous 2 weeks), and is not necessarily representative of chronic depression which requires successive depression measurements over time. For this reason, depression screening integrated into HIV services should occur at all or multiple visits for optimal risk identification and service provision. The small sample size also likely prevented the ability to associate depression with HIV and other morbidities, and the results are not generalizable beyond this sample. Finally, while the PHQ-9 is used extensively worldwide in both research and practice settings and has been validated for use in Peru, it is intended as a depression screening tool; a definitive depression diagnosis requires additional assessment^[57]^.

## CONCLUSIONS

People at risk for or living with HIV face an increased and synergistic risk of depression and poorer HIV-related health outcomes warranting the integration of depression care with HIV prevention and care services. While severe depression requires specialized care, moderate depression, which is associated with the greatest levels of HIV risk behaviors, could be treated by NSHWs using existing low-intensity depression interventions. LMICs could begin by integrating low-intensity depression interventions already in use with HIV prevention and treatment services.

## Data Availability

Data are available upon request.

## AUTHOR CONTRIBUTIONS

JTG, SRL, HS and BB conceived the data analysis, HS compiled the dataset and ZZ conducted performed the statistical data analysis. JTG wrote the first draft of the manuscript, which was reviewed and edited by all authors.

## ACKNOWLEDGMENTS

The authors express gratitude to Epicentro for provision of these data and their important work towards integration of depression and HIV care in Lima, Peru and to Caroline Muster for manuscript preparation support.

